# Using Random Forest feature importance results to predict zoonosis

**DOI:** 10.1101/2023.05.04.23289530

**Authors:** Roger Geertz Gonzalez

## Abstract

This study fills a gap in the literature regarding using machine learning techniques within the field of zoonoses. Instead of using linear and logistic inference modeling like in previous (Knowledge, Attitudes, and Practices (KAP) studies, this study incorporates Random Forest (RF) to identify important features that predict zoonotic diseases using survey and blood serology data. Using RF, we found that the most important features related to zoonoses were villages where households were 46 or larger and where participants owned many animals such as ducks, cats, and pigs. Compared to previous KAP studies in other countries where ethnicity, age, and education background were important features regarding knowledge, awareness, and practices relating to zoonoses, the KAP Cambodia case was different because these features were not found to be important.

## 1. Introduction

The outbreak of COVID-19 in 2020 demonstrated the importance of disease surveillance and its predictive ability of spillover events. Zoonotic spillover events occur when a reservoir animal population with a high concentration of pathogens comes into contact with humans and transmits the pathogens to humans which may or may not infect other humans. Although many zoonotic diseases like Ebola are well-known, a zoonotic outbreak can still occur like it did in 2013 in Guinea, Sierra Leone, and Liberia (Bird & Mazet, 2018). Disease surveillance works effectively when human capital and technical expertise along with sustainable funding, training programs, and engagement with national and international partners are crucial.

KAP surveys are central to disease surveillance programs because they represent zoonotic health related information about a specific population (Ul Haq et al., 2012). Knowledge questions in the KAP survey are asked to determine how aware the population is about specific health topics such as how diseases are transmitted. The Attitude section focuses on questions that determine how people think, feel, and act toward a specific health topic. The Practices part looks at what type of preventive measures people take regarding health issues (Ul Haq et al., 2012). For example, in their KAP study in Tanzania, Kiffner et al. (2019) found that the older respondents were aware of anthrax and how it spread more than younger respondents. They also found that males in certain districts were more knowledgeable about anthrax than females. Additionally, the Maasai and Arusha were less aware of brucellosis than other ethnicities. Older respondents had more knowledge of rabies in three districts. In Bhutan, respondents were unable to recognize rabies in cattle (Rinchen et al., 2019). They also found that respondents in the southern and eastern parts of Bhutan were not aware of preventive vaccinations for rabies in cattle. (Saylors et al., 2021) identified that those > 30 in Cameroon were most likely to eat rodents than those < 30. They also found that most respondents were unaware that bushmeat can spread zoonotic diseases. Respondents in Kazakhstan reported high risk activities for tick borne diseases such as: handling raw meat, shearing, herding, and slaughtering animals without wearing personal protective equipment (PPE) (Head et al., 2020).

Another important disease surveillance tool is rapid serological diagnostics of non-healthy carriers of disease, susceptible hosts where infection leads to pathogen replication and infection (Bird & Mazet, 2018). Serological analyses are important because they show presence of antibodies for specific diseases (Bird & Mazet, 2018).

Infectious disease modeling is essential for understanding and testing different public health strategies to prevent future epidemic outbreaks (Dattner & Huppert, 2018). The above studies used either linear or logistic regression for statistical inference modeling, or to determine the relationship between variables in their studies of infectious disease (Funk & King, 2020). For this study, we specifically focus on machine learning prediction techniques to predict positive and negative cases and to determine which features are the most important that might cause positive cases. There are recent examples where machine learning methods to predict infectious diseases were successful. For example, Han et al. (2015) used machine learning techniques to identify reservoir status with high accuracy and predicted new hyperreservoir (harboring 2 or more zoonotic pathogens). (Colubri et al. (2016) created a machine learning model to predict clinical outcomes in patients seropositive with Ebolavirus during the 2013-2016 West African epidemic.

For our Random Forest (RF) classifier and feature extractor machine learning model, we used antibody test results as the target variables. The viruses for this study originated from bats occurring on a global scale. They included paramyxoviruses and filoviruses. Paramyxoviruses like Ghanian bat henipavirus (GHV), Mojiang virus (MOJV), and Menangle virus (MENV) are spread to humans and livestock via pteropid bats (Halpin et al., 2011; (Mbu’U et al., 2019). GHV and MOJV are henipaviruses. GHV was isolated in Ghana in pigs and later Ghanian fruit bats were found to be infected to GHV (Mbu’U et al., 2019). MOJV was first identified after six miners died in a Mojiang cave in China (Rahalkar & Bahulikar, 2020). The virus was isolated from a rat sample in the mine. However, more recent analysis of the virus shows that it came from horseshoe bats in the Mojiang cave (Rahalkar & Bahulikar, 2020). MENV was first isolated in New South Wales in 1997 at a piggery (Wang & Anderson, 2019). Grey-headed flying foxes and black flying foxes were deemed to be the source of the MENV outbreak at the piggery.

Br et al. (2019) found that fruit bats had at least three filoviruses (Ebolavirus [EBOV], Bundibugyo virus [BDBV], and Sudan ebolavirus [SUDV]) in northern India that exhibited cross-reactivity and were present in bat hunters. (Letko et al., (2020), found that the bat-borne pig virus Swine acute diarrhea syndrome is responsible for killing thousands of pigs. In their study in Bangladesh, Chowdhury et al. (2014) found that henipavirus infected cattle, goats, and pigs. They also found that horses and cats had been infected by henipaviruses. Similarly, in this study, we found that important features for specific bat-borne viruses included cattle, pigs, cats, and pigs. Bombali virus (BOMBV) was discovered in 2018 in Sierra-Leone in free-tailed bats (Wang & Anderson, 2019). Because BOMBV was discovered exclusively in bats, it suggests that bats are likely reservoirs for ebolaviruses in general (Wang & Anderson, 2019).

The KAP Cambodia Survey “Knowledge” section (see Appendix A) has a True or False part including questions asking participants various questions related to the safe handling of wildlife and knowledge of zoonotic diseases. It also consists of open-ended questions regarding wildlife disease knowledge asking which diseases are transmitted by wild animals, what are some clinical signs of diseases transmitted by wild animals, and how we can protect ourselves from these diseases. In the report following the 2016 survey, most of the participants supported the conservation of wildlife and to a considerable extent understood the potential effects of zoonosis occurrence, as well as the proper practices to prevent harmful disease outbreaks as revealed in their responses to the following normative statements (International Development Centre, 2017). Some respondents were familiar with several procedures to protect themselves from wildlife disease infections, as well, including the maintenance of sanitary home environments. However, there were also some participants who were unaware of those procedures.

In the KAP Survey, the “Attitudes” section is a Likert scale section asking 7 specific questions about how participants feel about consuming, eating, and trading wildlife. Answers can be “acceptable,” “neutral,” or “totally unacceptable.” The “Practices” section contains 16 questions regarding the keeping of wildlife pets, how participants prepare wildlife for consumption, and how they trade wildlife. Answers throughout are a mix of “yes” and “no” questions, open ended questions, and Likert scale questions. Attitudes about eating wildlife in the 2016 survey were about evenly distributed between “totally acceptable”, “neutral”, and totally “unacceptable” responses (International Development Centre, 2017). Most had a neutral attitude about eating wildlife for medicinal purposes. Over half of the respondents stated that it was totally unacceptable to own wildlife. Most participants also stated that capturing wildlife was totally unacceptable.

The practices section consisted of 16 questions pertaining to consuming, owning, trading, working with wildlife as well as which actions, they take when they get sick related to eating and handling wildlife. More than half stated they owned livestock or owned a pet and most stated they engaged in high-risk behavior with wildlife contact (International Development Centre, 2017). More than half reported contact with wildlife using wild meat as food. Participants stated that they were the closest in proximity to mammals they used as pets, food consumption, or owning livestock. Wild pig, red muntjac, and rats were the most popular mammals eaten. Rats were popular with those on the boundary between Cambodia and Vietnam. The second highest category of animals eaten were birds such as spotted doves, sparrows, and cattle egrets. The third category of animals most often eaten were reptiles including snakes, turtles, and lizards. Participants also stated they had contact with insects such as crickets, worms, and bees. Most respondents stated they preferred their wildlife cooked thoroughly. Spotted doves and parrots were the most popular pets owned. About half reported taking their pets to veterinarians. However, most reported that they treated their diseased pets themselves. The most traded wildlife species were wild pigs, red mutjac, rabbits, spotted dove, parrot, green pigeon, and fish. In provinces, Stueng Treng, Preah Sihanuk, Rattanakiri, Siem Reap, Kampot, and Preah Vihear, traders primarily preferred hunting to obtain wild animals to be traded, while traders in other provinces obtained the animals to be traded not only by means of hunting, but by other means, as well, especially purchases from other hunters or other wholesale traders. Based on previous KAP survey results, scientists recommend refraining from touching sick domestic or wild animals; touching or eating wild bush meat; eating uncooked or fresh wild meat; capturing wild animals from forests; and engaging in the wildlife trade, as well as wearing protective clothing or materials when contacting wildlife and informing local authorities, veterinarians, and health officers of instances of zoonosis outbreaks.

The objective for this study was to incorporate RF feature extraction and classifier to identify the important variables from the KAP survey using seven target variables: Menangle virus (MENV), Bombali ebolavirus (BOMBV), Zaire ebolavirus (EBOV), Bundibugyo virus (BDBV), Sudan ebolavirus (SUDV), Mojiang virus (MOJV), and Ghanian bat henipavirus (GHV). Our research questions were: 1) How important are age, gender, education, and regional features regarding knowledge, attitudes, and practices of zoonoses in wildlife trade and how well do they predict zoonoses according to the serology data?; 2) How important is the type of food participants eat and trade regarding zoonoses in general?; 3) Are each of these features related more to each other and how are these features important that are different than ones pointed out in the literature above because they are related directly to Cambodia? Direct transmission from bats to animals to humans are the major causes of zoonotic infections as well as human to human transmission and thus, for this study, the RF important features included animals and human to human transmission features as part of the predictive RF model.

## 2. Materials and Methods

*The Knowledge, Attitudes, and Practices: Survey of Zoonoses in Wildlife Trade* (KAP) questionnaire was developed and administered to participants in Cambodia 2016, 2018, and 2019. 1,555 KAP surveys were administered by interviewers in 98 communes, 73 of 170 districts located in 21 of the country’s 24 provinces (see Figure 2.1) as well as Phnom Penh (Forestry Administration, 2017). Demographic questions included: Gender, age, marital status, ethnicity, religion, highest educational level, occupation, monthly income, and size of household. The survey consisted of 3 sections asking questions related to zoonoses and wildlife trade (Table 2.1).

**Figure 2.1.**
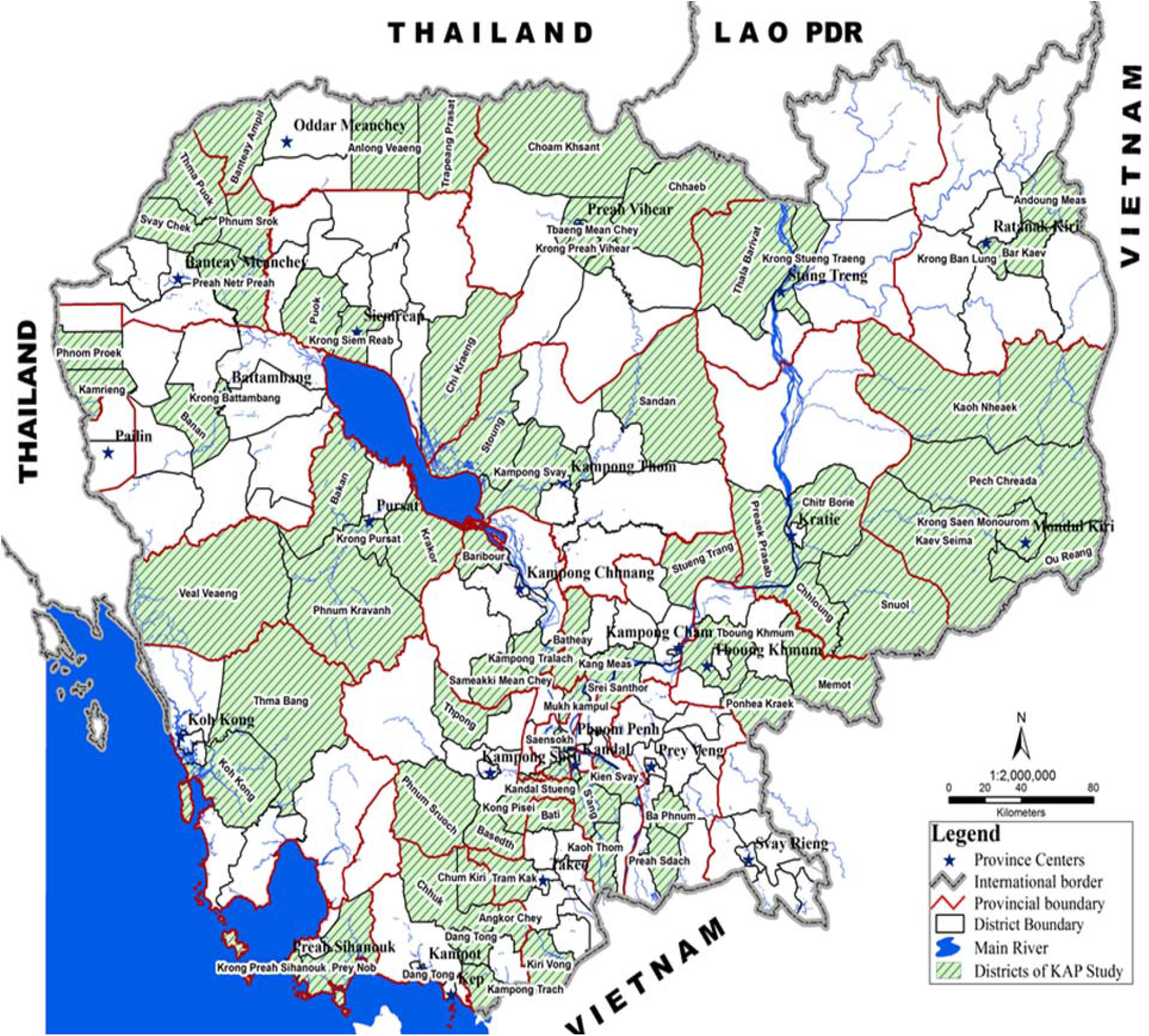
Cambodia map showing sample sites from KAP Survey from International Development Research Centre, 2017, Administrative map of Cambodia.

**Table 2.1.**
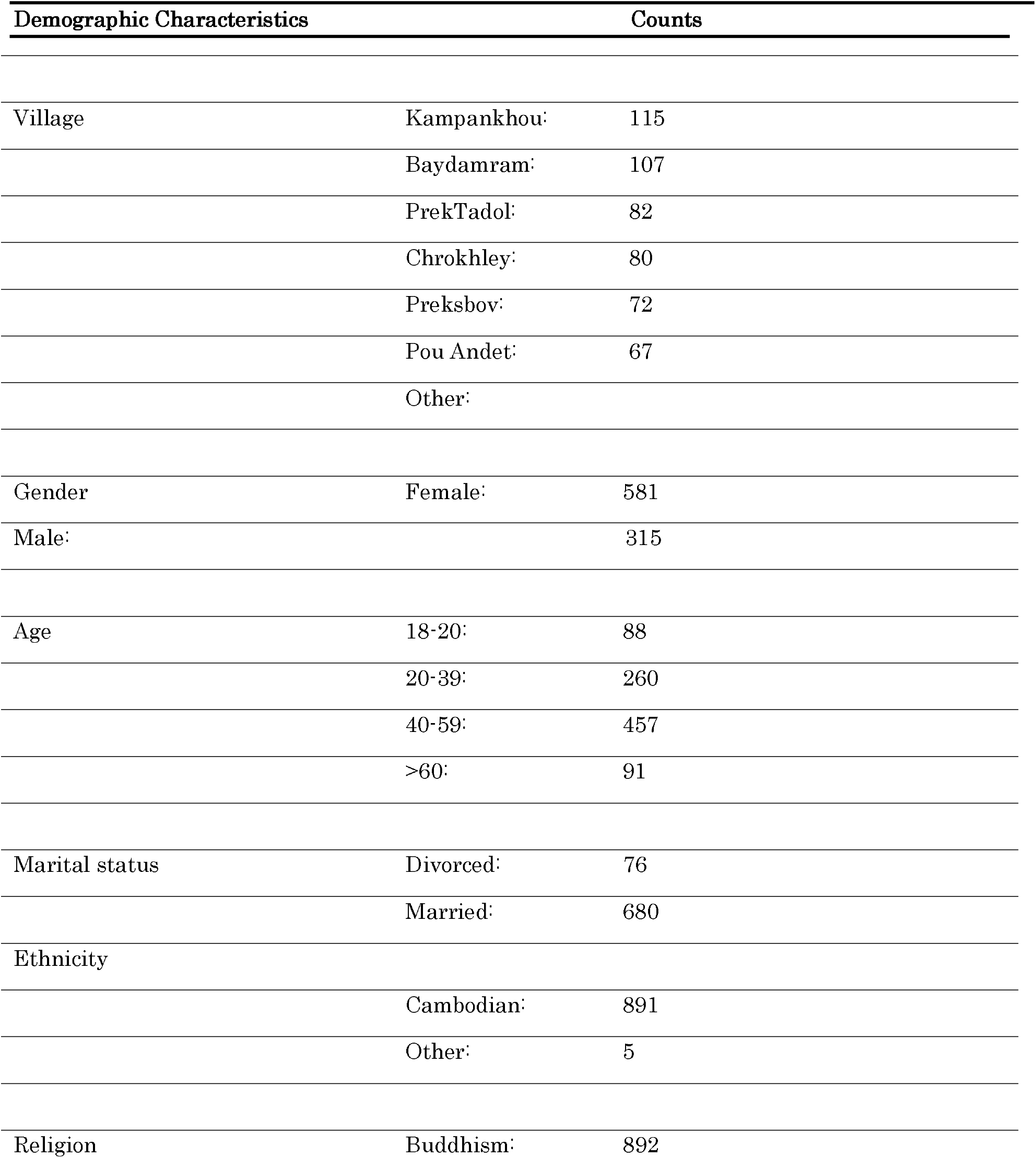
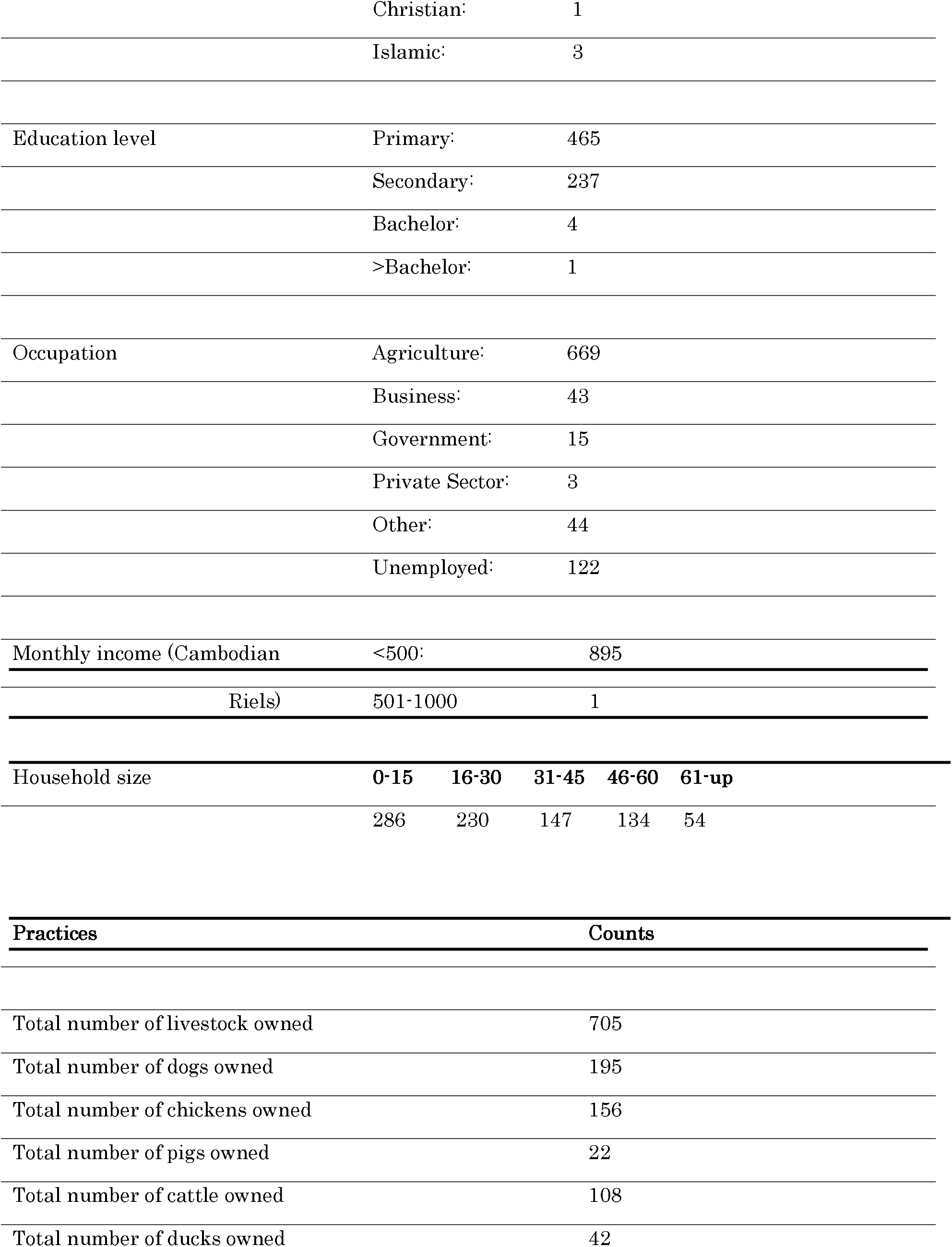
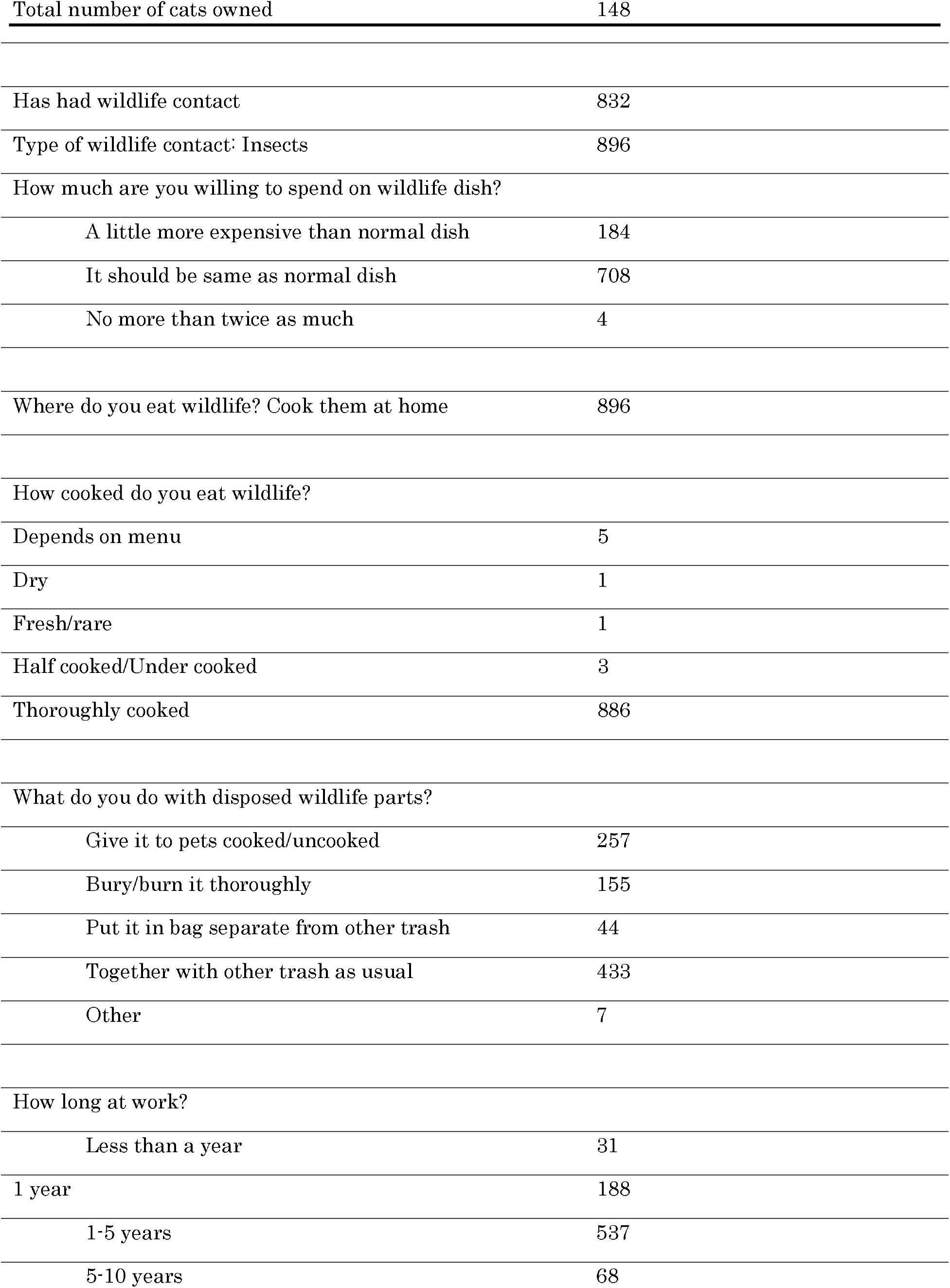
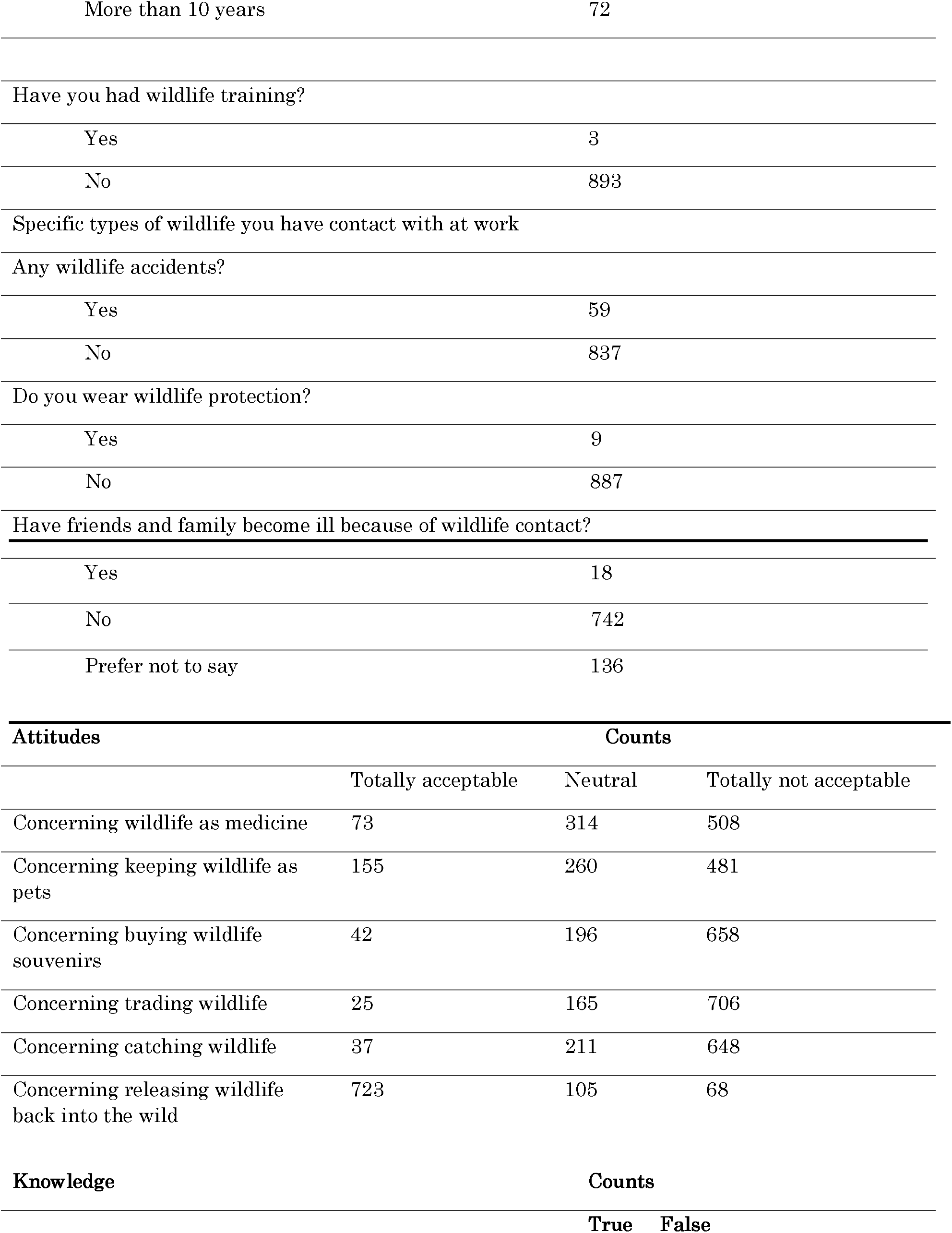
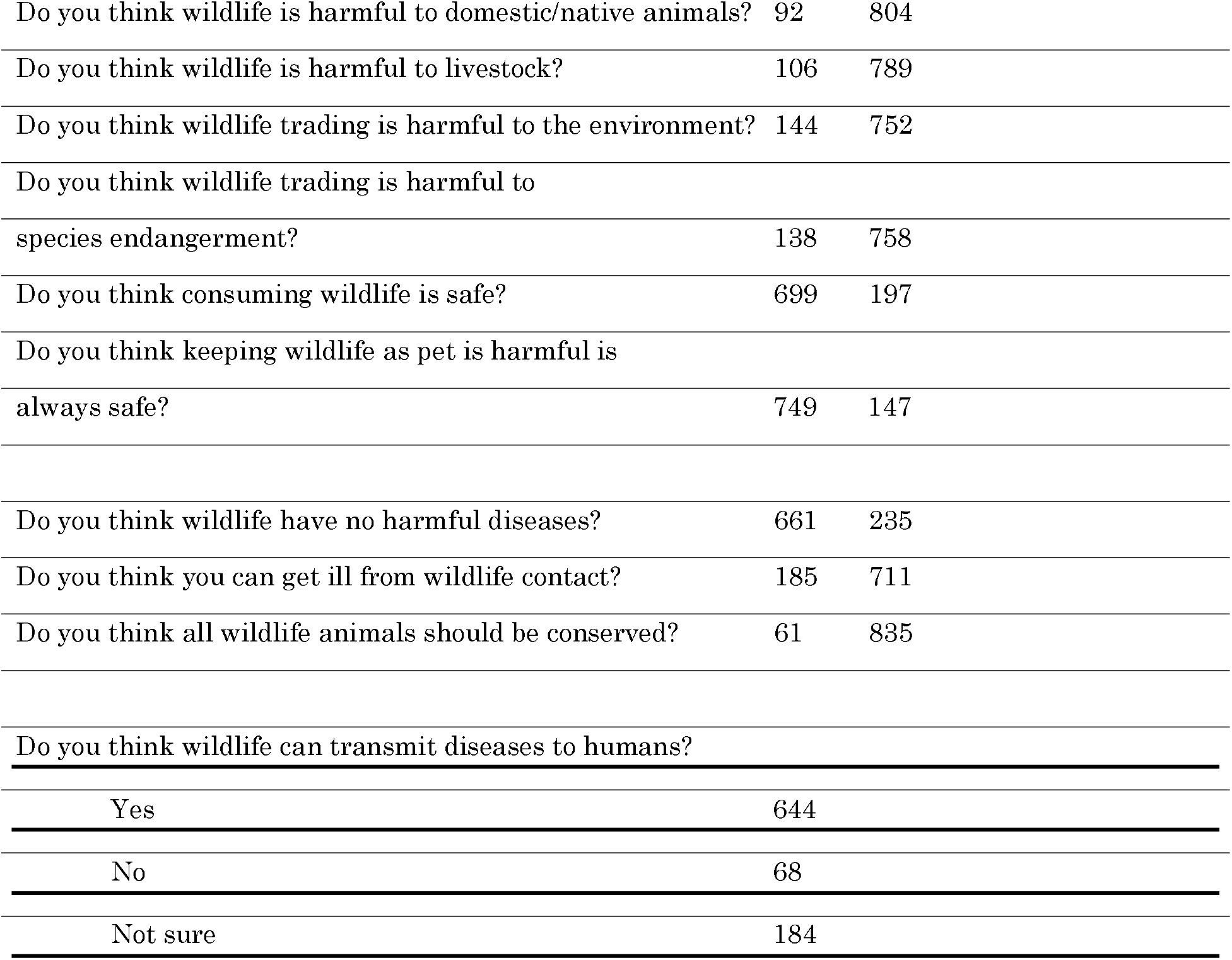
Feature Counts Used for Random Forests Classifier and Feature Extraction

| Demographic Characteristics |  | Counts |
| --- | --- | --- |
| Village | Kampankhoul | 115 |
|  | Baydamram | 107 |
|  | PrekTadol | 82 |
|  | Chrokhley | 80 |
|  | Preksbov | 72 |
|  | Pou Andet | 67 |
|  | Other |  |
| Gender | Female | 581 |
|  | Male | 315 |
| Age | 18-20 | 88 |
|  | 20-39 | 260 |
|  | 40-59 | 457 |
|  | >60 | 91 |
| Marital status | Divorced | 76 |
|  | Married | 680 |
| Ethnicity |  |  |
|  | Cambodian | 891 |
|  | Other | 5 |
| Religion | Buddhism | 892 |
|  | Christian: | 1 |
|  | Islamic: | 3 |
| Education level | Primary: | 465 |
|  | Secondary: | 237 |
|  | Bachelor: | 4 |
|  | >Bachelor: | 1 |
| Occupation | Agriculture: | 669 |
|  | Business: | 43 |
|  | Government: | 15 |
|  | Private Sector: | 3 |
|  | Other: | 44 |
|  | Unemployed: | 122 |
| Monthly income (Cambodian | <500: | 895 |
| Riels) | 501-1000 | 1 |

| Household size | 0-15 | 16-30 | 31-45 | 46-60 | 61-up |
| --- | --- | --- | --- | --- | --- |
|  | 286 | 230 | 147 | 134 | 54 |

| Practices | Counts |
| --- | --- |
| Total number of livestock owned | 705 |
| Total number of dogs owned | 195 |
| Total number of chickens owned | 156 |
| Total number of pigs owned | 22 |
| Total number of cattle owned | 108 |
| Total number of ducks owned | 42 |
| Total number of cats owned | 148 |
| Has had wildlife contact | 832 |
| Type of wildlife contact: Insects | 896 |
| How much are you willing to spend on wildlife dish? |  |
| A little more expensive than normal dish | 184 |
| It should be same as normal dish | 708 |
| No more than twice as much | 4 |
| Where do you eat wildlife? Cook them at home | 896 |
| How cooked do you eat wildlife? |  |
| Depends on menu | 5 |
| Dry | 1 |
| Fresh/rare | 1 |
| Half cooked/Under cooked | 3 |
| Thoroughly cooked | 886 |
| What do you do with disposed wildlife parts? |  |
| Give it to pets cooked/uncooked | 257 |
| Bury/burn it thoroughly | 155 |
| Put it in bag separate from other trash | 44 |
| Together with other trash as usual | 433 |
| Other | 7 |
| How long at work? |  |
| Less than a year | 31 |
| 1 year | 188 |
| 1-5 years | 537 |
| 5-10 years | 68 |
| More than 10 years | 72 |
| Have you had wildlife training? |  |
| Yes | 3 |
| No | 893 |
| Specific types of wildlife you have contact with at work |  |
| Any wildlife accidents? |  |
| Yes | 59 |
| No | 837 |
| Do you wear wildlife protection? |  |
| Yes | 9 |
| No | 887 |
| Have friends and family become ill because of wildlife contact? |  |
| Yes | 18 |
| No | 742 |
| Prefer not to say | 136 |

| Attitudes | Counts |  |  |
| --- | --- | --- | --- |
|  | Totally acceptable | Neutral | Totally not acceptable |
| Concerning wildlife as medicine | 73 | 314 | 508 |
| Concerning keeping wildlife as pets | 155 | 260 | 481 |
| Concerning buying wildlife souvenirs | 42 | 196 | 658 |
| Concerning trading wildlife | 25 | 165 | 706 |
| Concerning catching wildlife | 37 | 211 | 648 |
| Concerning releasing wildlife back into the wild | 723 | 105 | 68 |
| Knowledge | Counts |  |  |
|  | True | False |  |
| Do you think wildlife is harmful to domestic/native animals? | 92 | 804 |
| Do you think wildlife is harmful to livestock? | 106 | 789 |
| Do you think wildlife trading is harmful to the environment? | 144 | 752 |
| Do you think wildlife trading is harmful to |  |  |
| species endangerment? | 138 | 758 |
| Do you think consuming wildlife is safe? | 699 | 197 |
| Do you think keeping wildlife as pet is harmful is |  |  |
| always safe? | 749 | 147 |
| Do you think wildlife have no harmful diseases? | 661 | 235 |
| Do you think you can get ill from wildlife contact? | 185 | 711 |
| Do you think all wildlife animals should be conserved? | 61 | 835 |
| Do you think wildlife can transmit diseases to humans? |  |  |
| Yes | 644 |  |
| No | 68 |  |
| Not sure | 184 |  |

Previous KAP studies have used linear and logistic regression for statistical inference modeling. The major limitation of linear and logistic regression is that they assume a linear relationship between the dependent variable and the independent variables (Lantz, 2019). In complex real-life situations relating to infectious diseases including causes, effects, and transmission, the relationship between the dependent variable and independent variables might be non-linear and not an oversimplified linear or logistic regression model. Another limitation for linear and logistic regression is that the independent variables (features in machine learning) must be known ahead of time and tested and then retested to ensure the model works. (Yoo et al., 2012)

Machine learning algorithms can augment both linear and logistic regression for both regression and classification problems for complex, real-life problems such as disease prediction and features do not need to be known ahead of time and then retested to ensure the best fitting model. For example, RF has been used successfully in disease prediction when it comes to identifying important features.

We used RF to extract the most important features and then used these to predict the targets which were the specific viruses identified as positive through the collected blood samples. RF has a high predictive accuracy regarding disease prediction. Uddin et al. (2019) found that one of the most accurate supervised machine learning algorithms for disease prediction was RF. Kamal Alsheref & Hassan Gomaa, 2019) found that RF had a > 90% accuracy for predicting blood disease. Alam et al. (2019) used RF on 10 different disease datasets ranging from disease and blood pressure and found that RF was the best classifier and feature extractor out of 6 other classifier and feature extractors.

Blood samples (serology) were collected from participants. The serology data of 1,095 participants includes glycoprotein results for 17 zoonotic viruses. Median fluorescence intensity (MFI), or the output values of microsphere binding assays, are the intensity of a specific antibody to a specific disease on a continuous scale. In Figure 2.2 below, for example, MFI levels are shown in a box plot showing zero has the lowest number and beyond 8000 to show the level of intensity of the specific antibodies for each virus. EBOV for example in Figure 1 has a maximum cuttoff of around 1000 which shows that anything above 1000 for EBOV means that it is seropositive, or positive for that disease.

**Figure 2.2.**
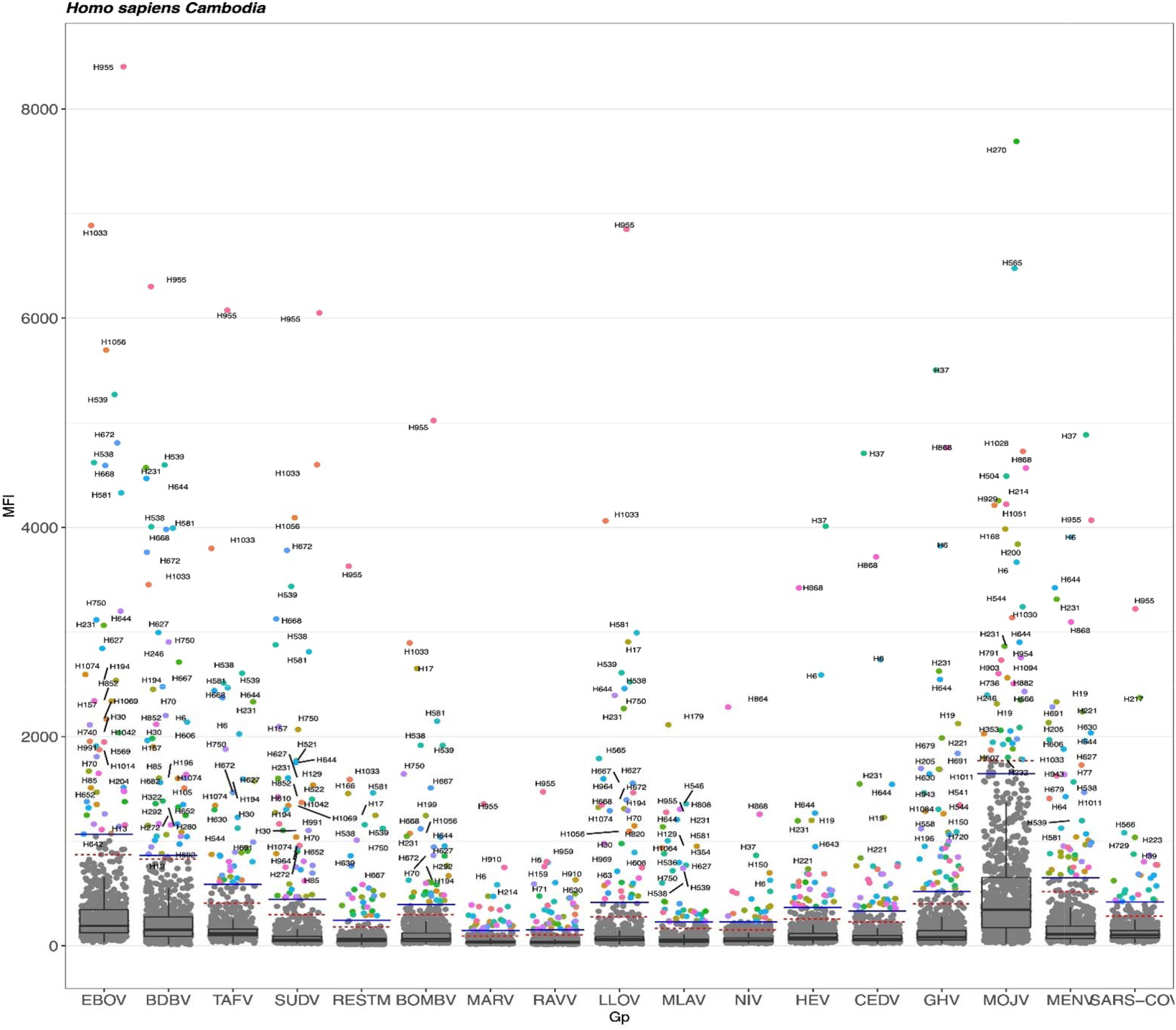
MFI Cutoffs *Note.* MFI 3 times above the arithmetic mean cutoffs are designated by a solid horizontal line. A dashed line indicates the log-normal cutoff. The 3 times the mean arithmetic cutoff was the highest positivity threshold used for this study.

Seropositivity cutoffs (if blood samples are considered positive or negative) were developed using 3-fold change above the arithmetic mean of the mock-adjusted scaled MFI data (shown as the solid straight line in the figure below) as well as fitted to a log-normal model (shown by dashed lines below) (unpublished data) (Figure 2.2). Viruses that exceeded the highest threshold, the 3-fold change above the arithmetic mean was considered seropositive (MENV, BOMBV, EBOV, BDBV, TAFV, SUDV, RAVV, LLOV, MLAV, MOJV, HEV, CEDV, and GHV).

Analyses for this study was done in R, version 4.1.0 (R Core Team, 2020). Missing imputation (MI) can reduce bias and improve efficiency for analysis of Missing at Random (MAR) data at any proportion of missingness (Madley-Dowd et al., 2019). For our data, missing values were treated as MAR, or the probability of missingness is independent of unobserved data because of the likelihood that some participants did not answer some questions about wildlife trade they might have found embarrassing or incriminating (Madley-Dowd et al., 2019). *missForest* was used to impute the missing values. It works well with continuous and categorical features (Stekhoven & Bühlmann, 2012). It uses the RF algorithm that averages over many unpruned classification or regression trees. The total missing values accounted for 9.81% of the total data. *missForest* iterated for a total of 9 times and replaced the missing values.

Data that show 0% to 4.7% positivity disease rates are difficult when creating training and testing for RF classifiers, because the classifiers assume that the test data is from the same distribution as the training data (Khalilia et al., 2011). Only diseases that had at least 4.7% positive cases or more were used to ensure the RF models had sufficient data for processing (Table 2.2). The *caret* package was specifically used for the RF classification and feature extraction (Kuhn, 2008). The data was divided into a training and test set, so the model could be evaluated on samples that were not used to build or tune the model and to provide an unbiased sense of model effectiveness (Kuhn & Johnson, 2016). The data was divided into 70% training and 30% testing.

**Table 2.2.**
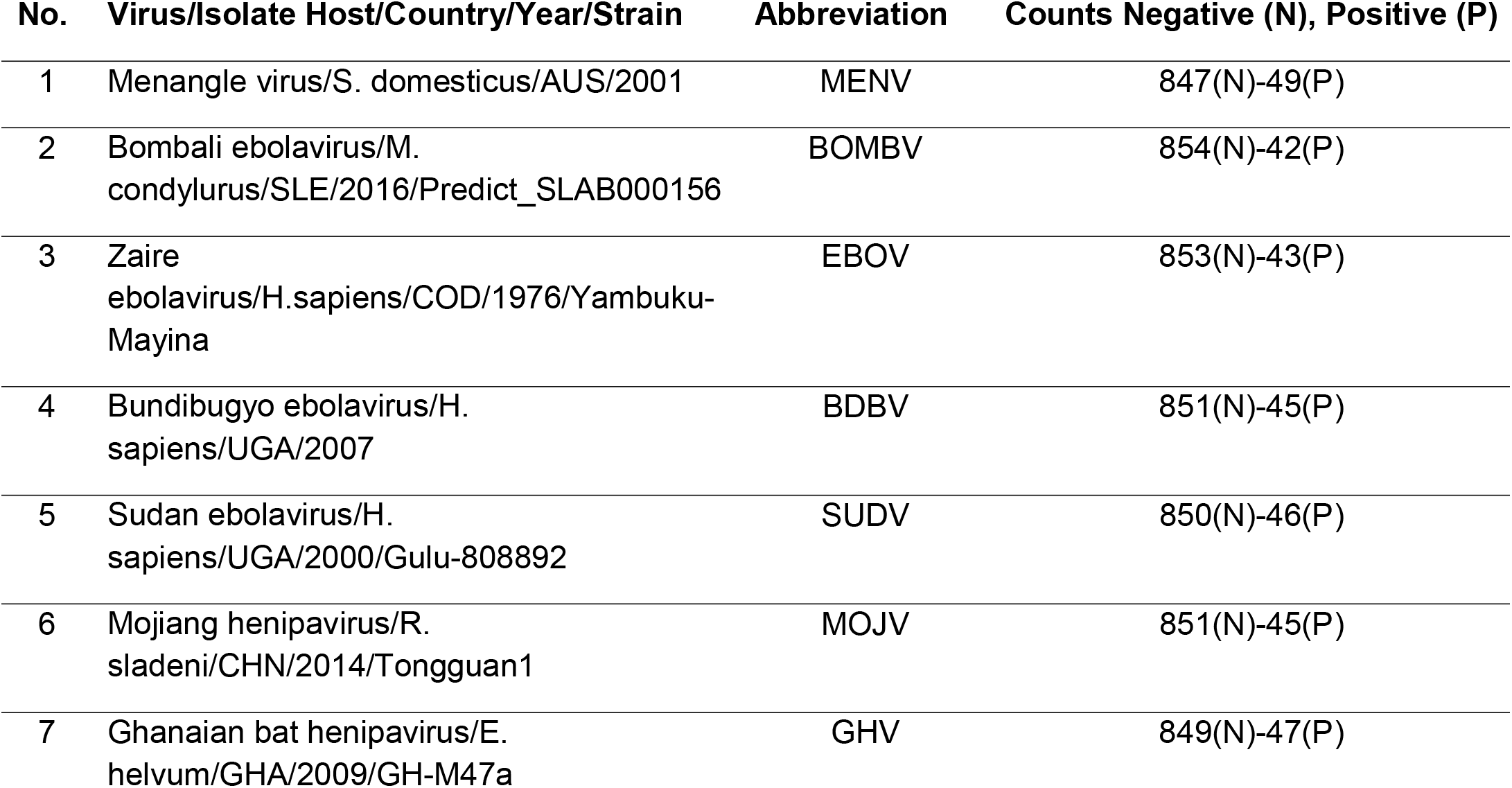
Specific Positive and Negative Counts of Zoonotic Diseases Used in Random Forests Analysis

The target variables were unbalanced where the majority were “negative.” The target variable classes were balanced using synthetic minority over-sampling technique (SMOTE) (Chawla et al., 2002; Kuhn & Johnson, 2016). SMOTE performs well when used for predictive RF classification (More & Rana, 2020); Marques et al., 2016). Quiroz et al. (2021) found that Covid-19 blood seropositivity improved positivity detection rates when SMOTE was included to machine learning classifiers including RF. We set the percent of the proportion over to 100, to keep a 1 to 1 proportion to balance the classes between negative and positive. 1000 trees were used because the linear combination of many independent learners reduces the variance of the overall ensemble relative to any individual learner in the ensemble (Kuhn and Johnson, 2016). Additionally, a ’for loop’ was created for the *mtry* function to identify how many random features in each split would produce the best results (Lantz, 2019). The ‘for loop’ identified between 3 and 8 random features per split with 99% accuracy. This is the only hypertuning function in the *caret* package for RF.

RF has a high predictive accuracy regarding disease prediction. Uddin et al. 2019) found that one of the most accurate supervised machine learning algorithms for disease prediction was RF. Kamal Alsheref & Hassan Gomaa, (2019) found that RF had a > 90% accuracy for predicting blood disease. Alam et al., (2019) used RF on 10 different disease datasets ranging from disease and blood pressure and found that RF was the best classifier and feature extractor out of 6 other classifier and feature extractors.

RF models produce two sets of feature importances: one based on the mean decrease of the gini index and the other based on permutation performance (Degenhardt et al., 2019). Mean decrease gini index feature importances have been shown to be biased (Degenhardt et al., 2019), we used permutation feature importances.

## 3. Results

The RF model accurately predicted the proportion of true positives and true negatives divided by the total number of predictions via the accuracy score (Table 3.1). The lowest accuracy score was for MOJV (0.94). The most important measures for classifying disease are accuracy, sensitivity, specificity, positive predictive values, negative predictive values, ROC, and AUC (Trevethan, 2017; Lantz, 2019). Accuracy specifically measures how often the model trained is correct, which is depicted by using the confusion matrix (Chen et al., 2020). Sensitivity measures the proportion of positive examples that were correctly classified (Lantz, 2019). Specificity measures the proportion of negative examples that were correctly classified. The positive predictive value is the proportion of positive examples that are truly positive. The negative predictive value is the proportion of negative examples that are truly negative. The ROC is commonly used to examine the tradeoff between the detection of true positives while avoiding the false positives. The AUC treats the ROC diagram as a two-dimensional square and measures the total area under the ROC curve. AUC scores are interpreted by the following: Outstanding=0.9 to 1.0, Excellent/Good=0.8 to 0.9, Acceptable/Fair=0.7 to 0.8, Poor=0.6 to 0.7, and No Discrimination=0.5 to 0.6. Below is a table (Table 3) showing the above RF performance measures.

**Table 3.1.** RF Classification Tree Statistics by Virus

| Disease | Accuracy | Sensitivity | Specificity | PPV | NPV | AUC |
| --- | --- | --- | --- | --- | --- | --- |
| MENV | **0.96** | **0.67** | **0.98** | **0.67** | **0.98** | **0.92** |
| BOMBV | **0.96** | **0.57** | **0.98** | **0.62** | **0.98** | **0.88** |
| EBOV | **0.98** | **0.77** | **0.99** | **0.83** | **0.99** | **0.93** |
| BDBV | **0.97** | **0.64** | **0.99** | **0.82** | **0.98** | **0.95** |
| SUDV | **0.98** | **0.71** | **1.00** | **0.91** | **0.98** | **0.81** |
| MOJV | **0.94** | **0.00** | **1.00** | NaN | **0.94** | **0.75** |
| GHV | **0.96** | **0.67** | **0.98** | **0.67** | **0.98** | **0.88** |

In positive disease classification, the most important metric is sensitivity or the true positive rate. RF sensitivity scores for, MENVV (0.67), BOMBV (0.57), EBOV (0.77), BDBV (0.64), SUDV (0.71), and GHV (0.67) were above 60%. MOJV was 0.00. Overall, the RF was able to correctly identify the proportion of negative cases that are truly negative via negative predictive values (NPV). The RF model had very high specificity scores (true negative rate). It had moderate to high positive predictive values (PPV) (probability of being truly positive) with one exception being MOJV with NaN, or unable to make the calculation. Additionally, according to the AUC scores, the RF models were able to distinguish between true positives (sensitivity) while avoiding false positives (specificity) except for MOJV which had a score of 0.75 (Figure 3.1).

**Figure 3.1.**
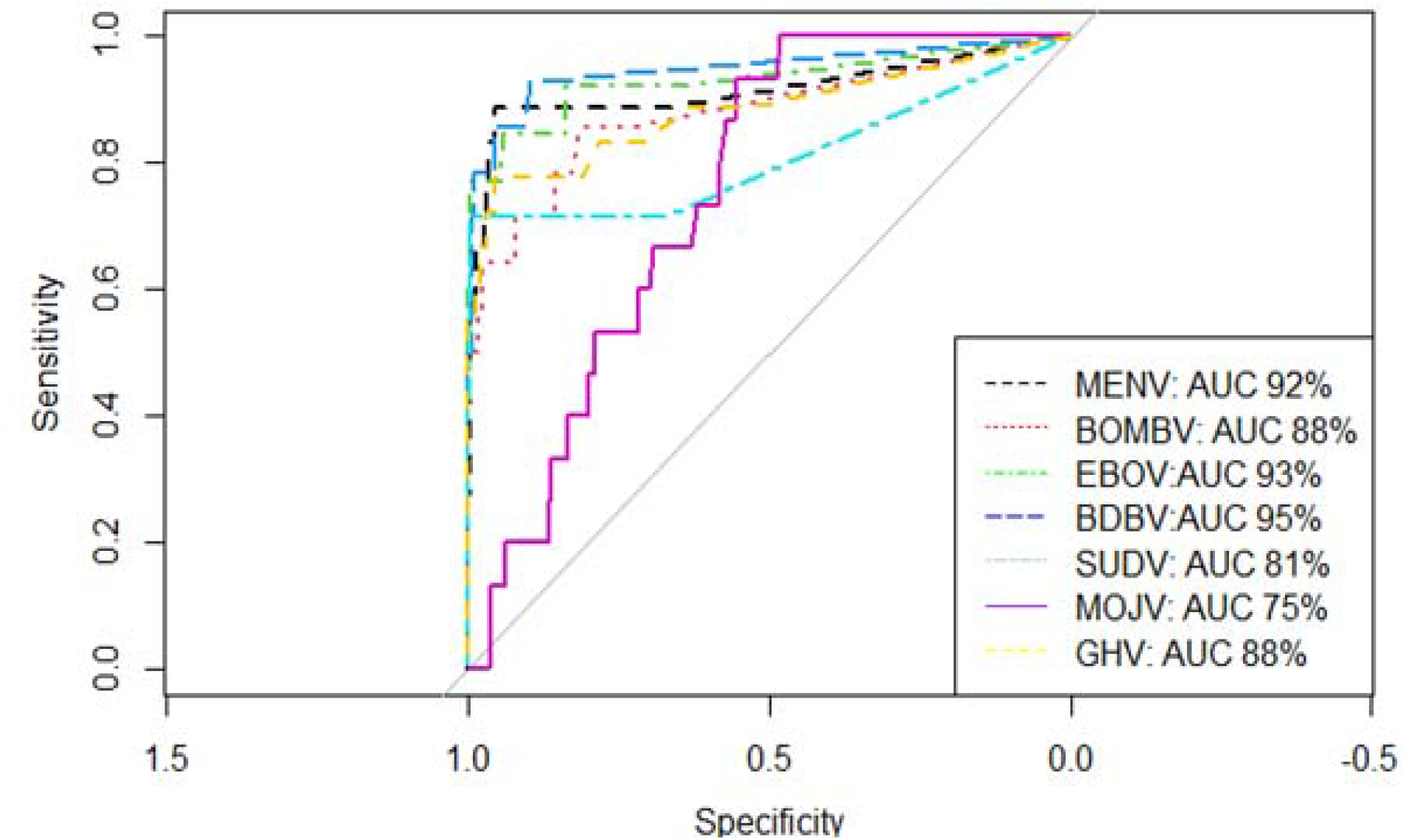
Random Forest Receiver Operator Curves for Each Disease *Note.* Receiver Operating Curves (ROC) curves for each disease (MENV, BOMV, EBOV, BDBV, SUDV, MOJV, and GHV) including individual Area Under Curve (AUC) percentages.

Important features for MENV were: GHV, village, and RAVV (Figure 3.2). For BOMBV, important features included: RESTm, LLOV, and village (Figure 3.2). The important features for EBOV, were SUDV, TAFV, number of pigs owned, and BDBV (Figure 3.2). The important features for BDBV, were LLOV, TAFV, EBOV, village, SUDV and MENV (Figure 3.2). The most important features for SUDV are EBOV, LLOV, TAFV, BDBV, cat, and pig (Figure 3.2). For MOJV, important features were pig ownership, duck ownership, village, cat ownership, and household size between 46 and 60 (Figure 3.2). Important features for GHV included: MENV, CEDV, and HEV (Figure 3.2).

**Figure 3.2.**
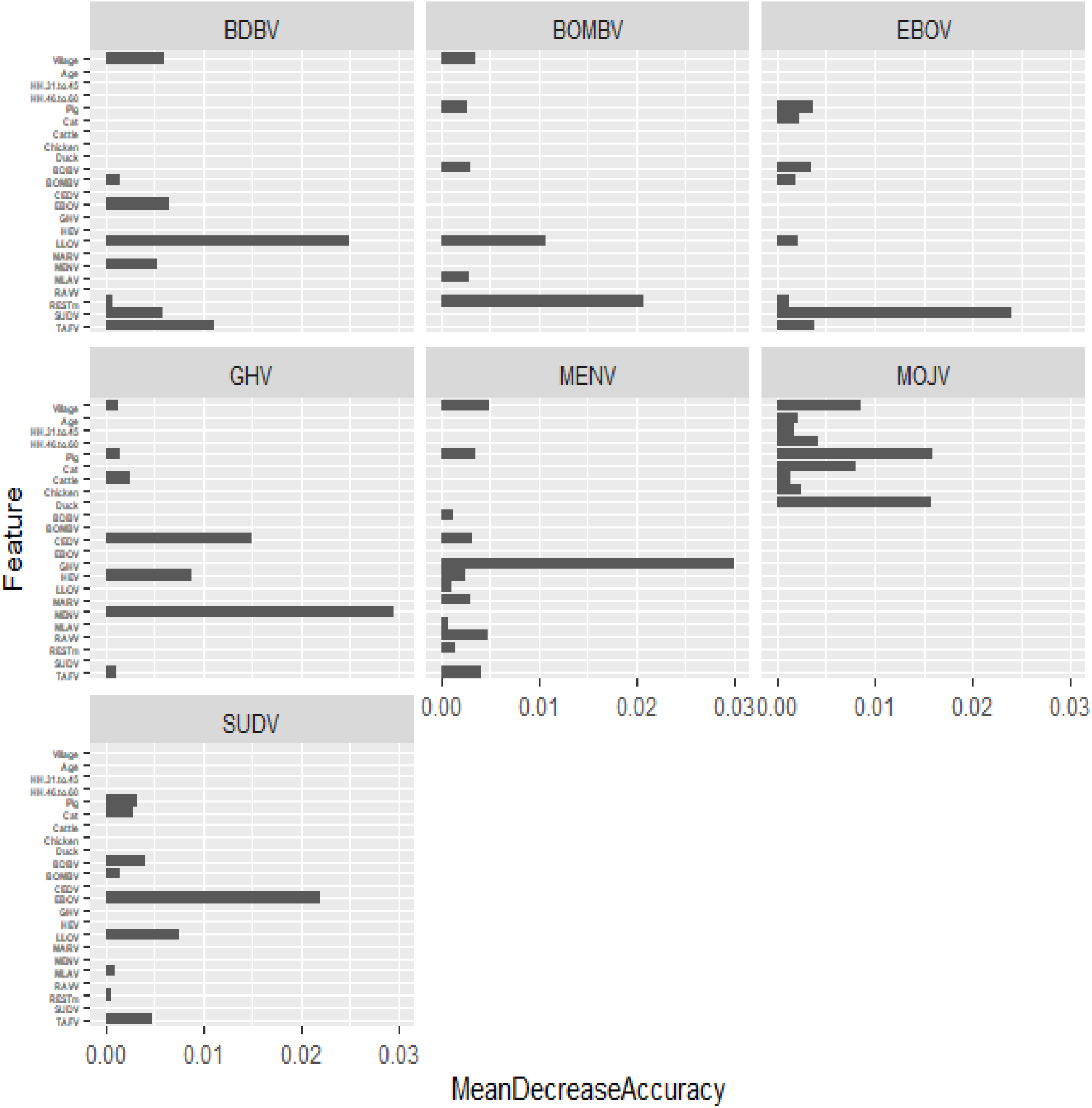
Feature importances for each virus

Village was important for the following diseases that were positive: MENV, BOMBV, BDBV, and MOJV (Figure 3.3). Chrokhley, Baydamram, Borbous, and Thmey were the villages where MENV had the most positive cases. Chrokhley, Baydamram, Borbous, Pou Andet, and Khouv had the most BOMBV positive cases. The most BDBV positive cases found were in Chrokhley, Baydamram, Borbous, and Pou Andet. Chrokhley, Baydamram, Borbous, Kamponghou, Prek Tadol, Thmey, and Thouv had the most MOJV positive cases. Presence of ducks was an important feature for MOJV (Figure 3.4). The highest number of duck ownership was found in Kampongkhou, Baydamram, Preksbov, and PrekTadol. Another bat-borne virus, Nipah virus has been found to infect cats (Glennon et al., 2018). For this study, cat was an important feature for SUDV and MOJV (Figure 3.4). The largest amount of cat ownership occurred in Kampongkhou, Baydamram, Preksbov, PrekTadol, and Chrokhley. Pigs were important features for EBOV, SUDV, and MOJV (Figure 3.4). Kampongkhou, Baydamram, Preksbov, PrekTadol, Borbous, and Andoungreang had the largest number of pig ownership. Number of households was an important feature for MOJV (Figure 3.5). Kampongkhou, Chrokhley, Baydamram, PouAndet. Preksbov, PrekTadol had the largest number of households between 46 and 60 people.

**Figure 3.3.**
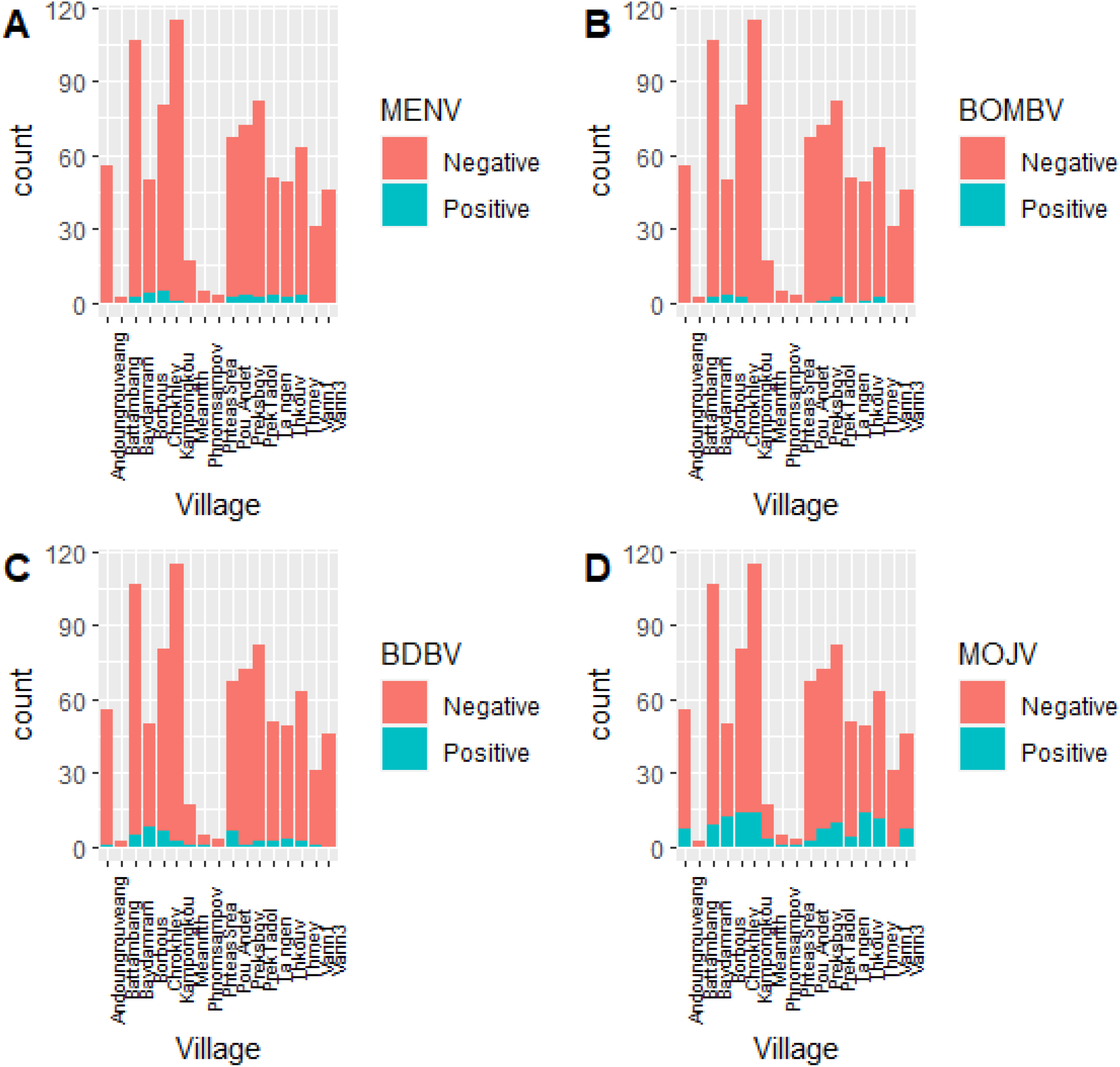
Disease counts in each village: A) MENV, B) BOMBV, C) BDBV, D) MOJV

**Figure 3.4.**
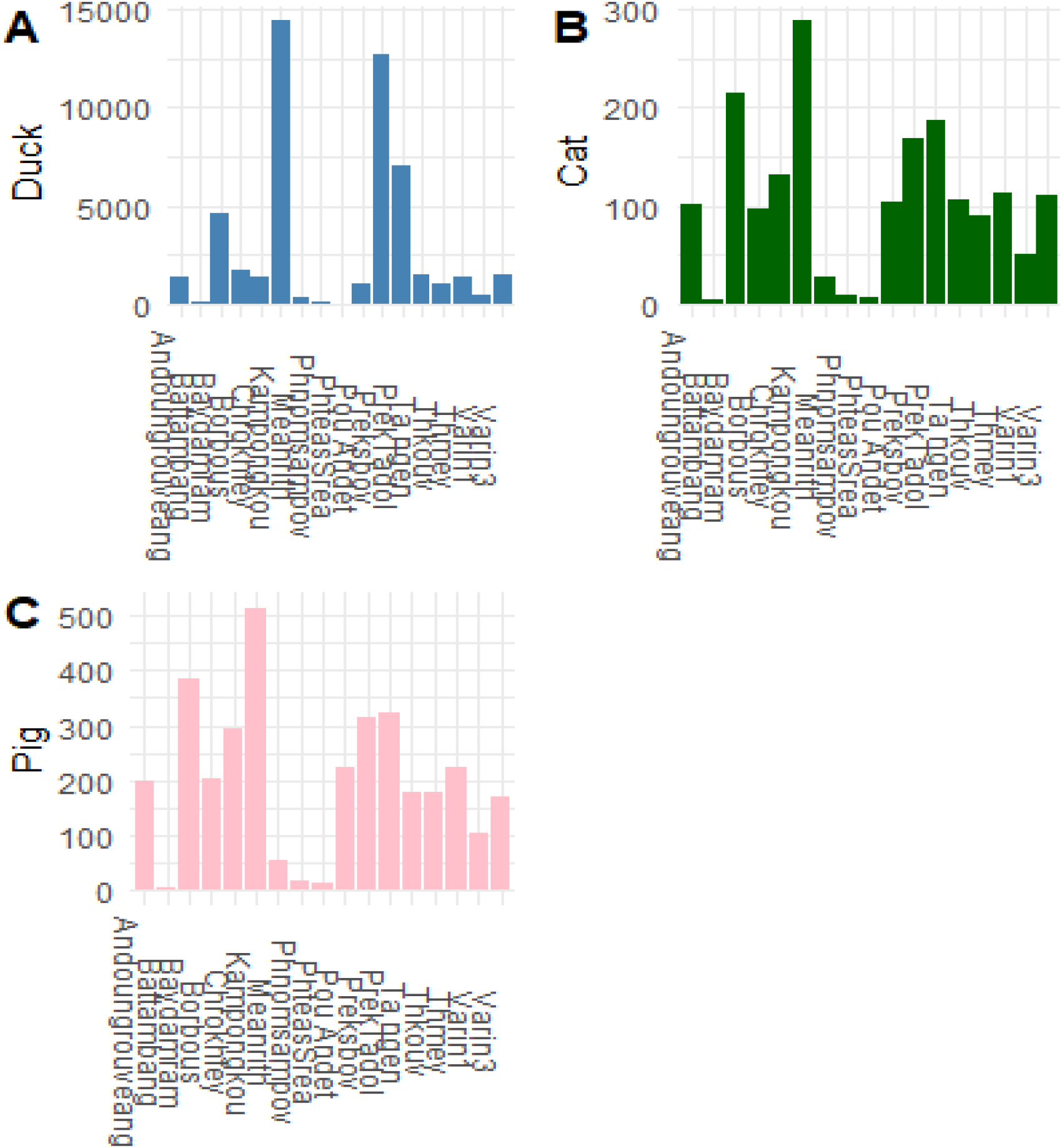
Animal counts by village: A-Duck, B-Cat, C-Pig

**Figure 3.5.**
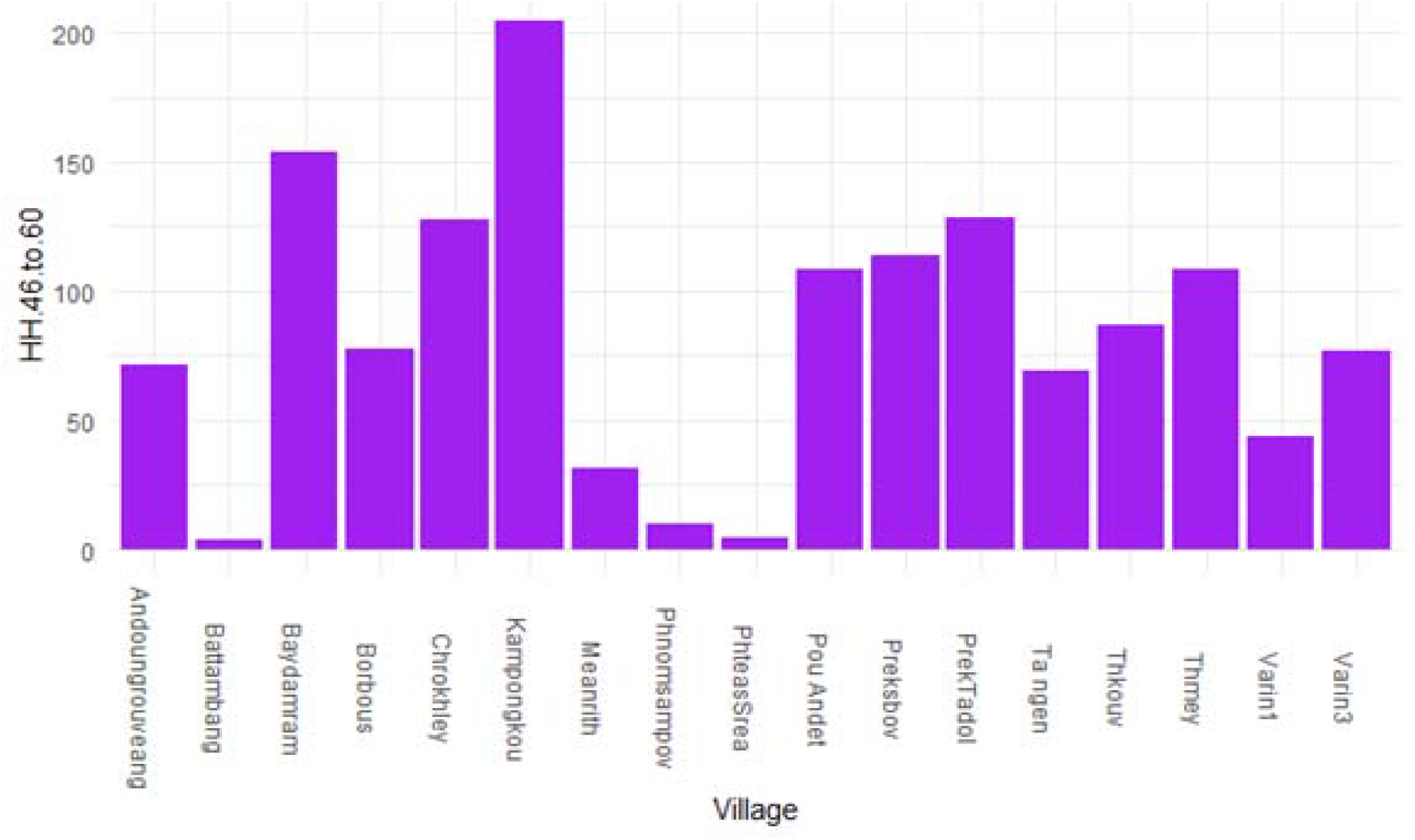
Number of Households 46 to 60 By Village

Since bats can transfer their infectious diseases to animals and then to humans (Mbu’U et al., 2019; Wang & Anderson, 2019), further research needs to be done to verify whether these animals in these specific villages in Cambodia are sick to then provide safety and disease spread mitigation efforts.

A connection between GHV and the type of pneumonia was also discovered by Rahalkar & Bahulikar (2020). They state that there is a possibility that it might be related to SARS-CoV-2. Pig is the most important feature and researchers highlight how bats can transmit diseases to pigs which then transmit them to humans (Mbu’U et al., 2019; Wang & Anderson, 2019). Pigs, ducks, and cats are predominant in Kampongkou (Figure 3.4).

Except for MOJV, the other 6 viruses had other viruses as feature importances. These could be due to cross-reactivity where antibody testing for one specific disease is like other diseases (Schuh et al., 2019).

## DISCUSSION

Our research questions were: 1) How important are age, gender, education, and regional features regarding knowledge, attitudes, and practices of zoonoses in wildlife trade and how well do they predict zoonoses according to the serology data?; 2) How important is the type of food participants eat and trade regarding zoonoses in general?; 3) Are each of these features related more to each other and how well are they related; and; 4) Are there features that are important that are different than ones pointed out in the literature above because they are related directly to Cambodia? When it comes to the first question, age, gender, and education were not important features. However, we did find that several villages had high seropositivity rates for MENV, BDBV, BOMBDV, and MOJV. These included Chrokhley, Baydamram, Borbous, Thmey, and PrekTadol. Regarding question two, eating specific foods was not found to be important. However, having ducks, pigs, and cats was found to be important since participants might trade these animals with each other. This could be the reason many participants showed that they were exposed to multiple diseases. Additionally, the same villages above along with Preksbov and Andoungreang were found to have the highest amount of these animals. Regarding question three, features that were related were having lots of ducks, pigs, cats, and households 46-60 in specific villages. Kampongkhou, Chrokhley, Baydamram, PouAndet, Preksbov, and PrekTadol had the highest number of households 46-60. For the last question, we found that Cambodia is unique compared to previous KAP studies because age and gender were not important features.

Previous KAP surveys use linear and logistic regression. The major limitation to linear and logistic regression is that they assume a linear relationship between the dependent variable and the independent variables (Lantz, 2019). In complex real-life situations relating to infectious diseases including causes, effects, and transmission, the relationship between the dependent variable and independent variables might be non-linear and not an oversimplified linear or logistic regression model. Another limitation for linear and logistic regression is that the independent variables (features in machine learning) must be known ahead of time and tested and then retested to ensure the model works. Machine learning algorithms like RF can replace both linear and logistic regression for both regression and classification problems for complex, real-life problems such as disease prediction and features do not need to be known ahead of time and then retested to ensure the best fitting model. Our RF model was able to find important features that successfully predict the likelihood of seropositivity.

When considering RF feature importances, disease surveillance should focus on cross-reactivity with other viruses, and household ownership of cats, ducks, and pigs. Br et al., (2019) found that fruit bats had at least three filoviruses (EBOV, BDBV, SUDV) in northern India that showed cross-reactivity and that were present in bat hunters. Swine acute diarrhea syndrome is a bat-borne virus that was responsible for killing thousands of pigs (Letko et al., 2020). Spillover of bat-borne viruses is caused in two ways: direct contact with bats or via intermediate hosts (Letko et al., 2020). Nipah virus has been rising in Malaysia via pigs. In Bangladesh, henipavirus infected cattle, goats, and pigs (Chowdhury et al., 2014). They also found that horses and cats had been infected by henipaviruses. Similarly, we found that important features for bat-borne viruses included cattle, pigs, and cats.

Since these viruses are similarly spread from bats to animals and humans, zoonoses disease experts should also be looking if these viruses might transmit together in specific combinations. Wang & Anderson (2019) point out that most zoonotic diseases like Ebolavirus, Mojiang virus and henipaviruses all come directly from bats. Thus, it would not be surprising then to find individuals that showed exposure to multiple zoonoses. When it comes to animal ownership, having a pig, duck and cat is related to all the viruses above.

Having households larger than 46 has an impact also on having the above viruses. Ebola viruses are most likely transmitted by bodily fluids between people (Judson et al., 2015). Large households could be contributing to virus transmission, in the Cambodia case according to the RF predictions. Perhaps owning pigs, ducks and cats means that people are in closer contact with each other since they are working together to improve their livelihoods by raising these animals for food and for selling and thus, they transmit these viruses to each other by working closely together (Wang & Anderson, 2019).

Bausch & Schwarz, (2014) demonstrates that economic survival of local villagers might blunt preventive efforts. For example, in Preksbov, there a lot of ducks owned and perhaps this means that economic survival depends on owning ducks. In their survey studying household practices and connections to zoonoses in Cambodia, Osbjer et al. (2015) found that poultry like ducks and livestock like cattle were used specifically for selling. According to Suttie et al. (2018), in 2013, 25 million chickens and 3.3 million ducks were either sold or slaughtered for sell or consumption in Cambodia. 22% of chickens and 18% of ducks there died from illness. In Preksbov, higher positive rates for EBOV may be due to interactions between duck owners, other people and other animals since EBOV is transmitted by bats to animals and humans (Osbjer et al., 2015). Additionally, Borbous and Thmey share 10 cases each where they have EBOV and have four other additional diseases. Therefore, it is important to closer examine these two villages and see how household size and ownership of pigs and cats and their interaction with bats might be infecting villagers with multiple diseases.

RF models are an effective tool providing good to excellent prediction performance when combining KAP surveys and serological data. Identification of positive cases which would be of more interest to infectious disease experts, RF does an excellent job on sensitivity, positively predicted values (PPV), and area under the curve (AUC) measures. RF should be included in any disease surveillance project that includes KAP surveys.

Disease mitigation for bat-borne viruses may include: providing villagers and local health experts with enough personal protective equipment (PPE) that would prevent transmission of disease such as masks and gloves; information campaigns should include educational programs that teach local villagers how best to protect themselves as livestock owners such as washing hands frequently and having local veterinarians frequently test livestock to ensure they are healthy; have local and international wildlife biologists track exactly where Cambodian villagers are interacting with bats and identify these so that these bats are tested and so that diseases are tracked; and finally, Cambodian villagers should be tested more frequently for multiple diseases instead of one at a time since in this study, it shows that multiple villagers have multiple diseases at once (Wang & Anderson, 2019; Glennon et al., 2018; Halpin et al., 2011). Wang & Anderson, 2019) state that many zoonotic diseases like Ebolavirus, Mojiang virus, and henipaviruses come directly from bats and then infect animals like livestock that in turn infect humans.

## Data Availability

All data produced in the present work are contained in the manuscript.

